# HematoNet: Expert Level Classification of Bone Marrow Cytology Morphology in Hematological Malignancy with Deep Learning

**DOI:** 10.1101/2022.04.30.22274525

**Authors:** Satvik Tripathi, Alisha Isabelle Augustin, Rithvik Sukumaran, Suhani Dheer, Edward Kim

## Abstract

There have been few efforts made to automate the cytomorphological categorization of bone marrow cells. For bone marrow cell categorization, deep-learning algorithms have been limited to a small number of samples or disease classifications. In this paper, we proposed a pipeline to classify the bone marrow cells despite these limitations. Data augmentation was used throughout the data to resolve any class imbalances. Then, random transformations such as rotating between 0° to 90°, zooming in/out, flipping horizontally and/or vertically, and translating were performed. The model used in the pipeline was a CoAtNet and that was compared with two baseline models, EfficientNetV2 and ResNext50. We then analyzed the CoAtNet model using SmoothGrad and Grad-CAM, two recently developed algorithms that have been shown to meet the fundamental requirements for explainability methods. After evaluating all three models’ performance for each of the distinct morphological classes, the proposed CoAtNet model was able to outperform the EfficientNetV2 and ResNext50 models due to its attention network property that increased the learning curve for the algorithm which was represented using a precision-recall curve.

## 1 Introduction

The human-based examination and characterization of bone marrow (BM) cells is one of the most important yet time expensive procedures in cancerous and non-cancerous hematological conditions [1, 2, 3, 4].

The cytomorphologic examination is still a critical initial step in the diagnosis of many intra- and extramedullary illnesses even though numerous advanced procedures like cytogenetics, immunophenotyping, and molecular genetics are now accessible [5, 6]. The function of BM cytology, which was created in the 19th century, is still very significant because of its relatively rapid results and extensive availability [7, 8]. Microscopic inspection and single-cell morphology categorization are still the primary responsibility of human clinicians due to the difficulty in automating this process. In certain circumstances, such as those involving ambiguous BM smears, the process of manually evaluating the specimens may be arduous and time-consuming [9, 10]. It has been observed that examiner classifications are prone to significant inter-and intra variability, which means that the number of high-quality cytological exams is constrained since subject matter experts are hard to come by [11, 12, 13, 14, 15].

It is also challenging to integrate this procedure with other diagnostic methods that provide more quantitative data since the analysis of individual cell morphologies is qualitative by nature. Few efforts have been made to automate the cytomorphological categorization of BM cells. Hand-crafted single-cell characteristics extracted from digital pictures are often used to categorize cells. Furthermore, most prior research on automated cytomorphologic classification focused on the classification of physiological cell types or peripheral blood smears, restricting their applicability to the classification of leukocytes in the BM for the diagnosis of hematological malignancies [16, 17, 18, 19, 20, 21, 5, 22]. For BM cell categorization, deep-learning algorithms have been limited to a small number of samples or disease classifications and/or have not made the related data accessible [23, 24, 25, 26, 27, 28, 29].

Convolutional neural networks have helped to significantly increase the accuracy of computer vision classification tasks in the last few years [30, 31, 32, 33, 34, 35]. These methods has also been used to identify multiple types of cancer, predict progression of tumor, and classify various types of skin diseases [36, 37, 38, 39, 40, 41]. Because of this, the effective use of CNNs for image classification depends on the availability of a substantial quantity of image data and high-quality annotation, which may be difficult to achieve because of the cost associated in collecting labels by medical specialists [42, 43, 44, 45]. Human examiners are required to supply the ground truth labels for network training and assessment in instances like cytomorphologic inspection of BM, when there is no underlying technological gold standard.

## 2 Related Work

### 2.1 Deep Learning Approaches

Deep Learning (DL) is an area of machine learning that uses artificial neural networks to model the brain’s structure and function. Deep learning powers numerous artificial intelligence (AI) apps and companies that automate analytical and physical processes. For cancer detection, Oncologists have been using Deep Learning methods to improve patient diagnosis, prognosis, and therapy selection by combining genomic, transcriptomic, and histopathological data. The purpose of DL is to develop decision-making tools to aid cancer researchers in their studies and health professionals in the clinical care of cancer patients [46]. In another case, Tayebi et al. worked on building an end-to-end deep learning-based model for automated bone marrow cytology [47]. A computerized full slide picture of the patient’s blood was used to quickly and automatically determine areas appropriate for cytology, which was followed by the identification and classification of all of the blood cells inside those areas. Cell type diversity in bone marrow is quantified by using the Histogram of Cell Types (HCT), a new visual depiction that serves as a cytological “patient fingerprint.” A great degree of precision was achieved in the method’s area detection [48].

### 2.2 Hematological Malignancy

In a similar research conducted on leukemia cancer, the study compared two leukemia detection methods.The first method was a genomic sequencing method and the second was multi-class classification model. Both employed a Convolutional neural network (CNN) as network design and also used three-way cross-validation to separate their datasets. The findings indicated that the genomic model performed better, with 98 percent accuracy in predicting values, whereas the Multi-class classification model has an accuracy of 81 percent. On the other hand, another research looked at the clinical usefulness of an array-based genome-wide screen in leukemia, as well as the technological obstacles and an interpretive procedure [13].

### 2.3 Bone Morrow Morphology

Another study by focused on developing an accurate bone marrow cell identification technique for quantitative analysis. The YOLOv5 network, trained by minimising a new loss function, was used in this study to offer a bone marrow cell identification technique. The suggested new loss function was based on a classification algorithm for detecting bone marrow cells. As per the results, the proposed loss function was beneficial in improving the algorithm’s efficiency, and the proposed bone marrow cell identification algorithm outperformed other cell detection techniques [49] Another research evaluated how useful flow cytometry, karyotype, and a fluorescence in situ hybridization (FISH) panel are in detecting myelodysplastic syndrome in children (MDS). The study concluded that flow cytometry and MDS FISH may be used in conjunction with morphological examination and karyotype to discover anomalies in specific cases [50].

## 3 Methods

### 3.1 Dataset

The dataset, acquired by Matek et al., contains 171,375 images from a cohort of 945 patients diagnosed with various hematological diseases at MLL Munich Leukemia Laboratory [51]. The minimum patient age was 18.1 years, and the maximum was 92.2 years. The average patient age, based off of the median, was 69.3 years. The mean age was 65.6 years.

Images of bone marrow smears were stained using May-Grünwald-Giemsa/Pappenheim staining. A brightfield microscope with 40x magnification and oil immersion were used to acquire the images. The original images were 2452×2056 in size. After individual cells were annotated into 21 different classes by morphologists, 250×250 square regions were extracted from the original images, each region containing one annotated cell.

Figure 1 indicates the classes of each image, as well as an example image of that class and the number of images from the original dataset in that class. The distribution of classes in the dataset can be seen in Figure 2.

**Figure 1:**
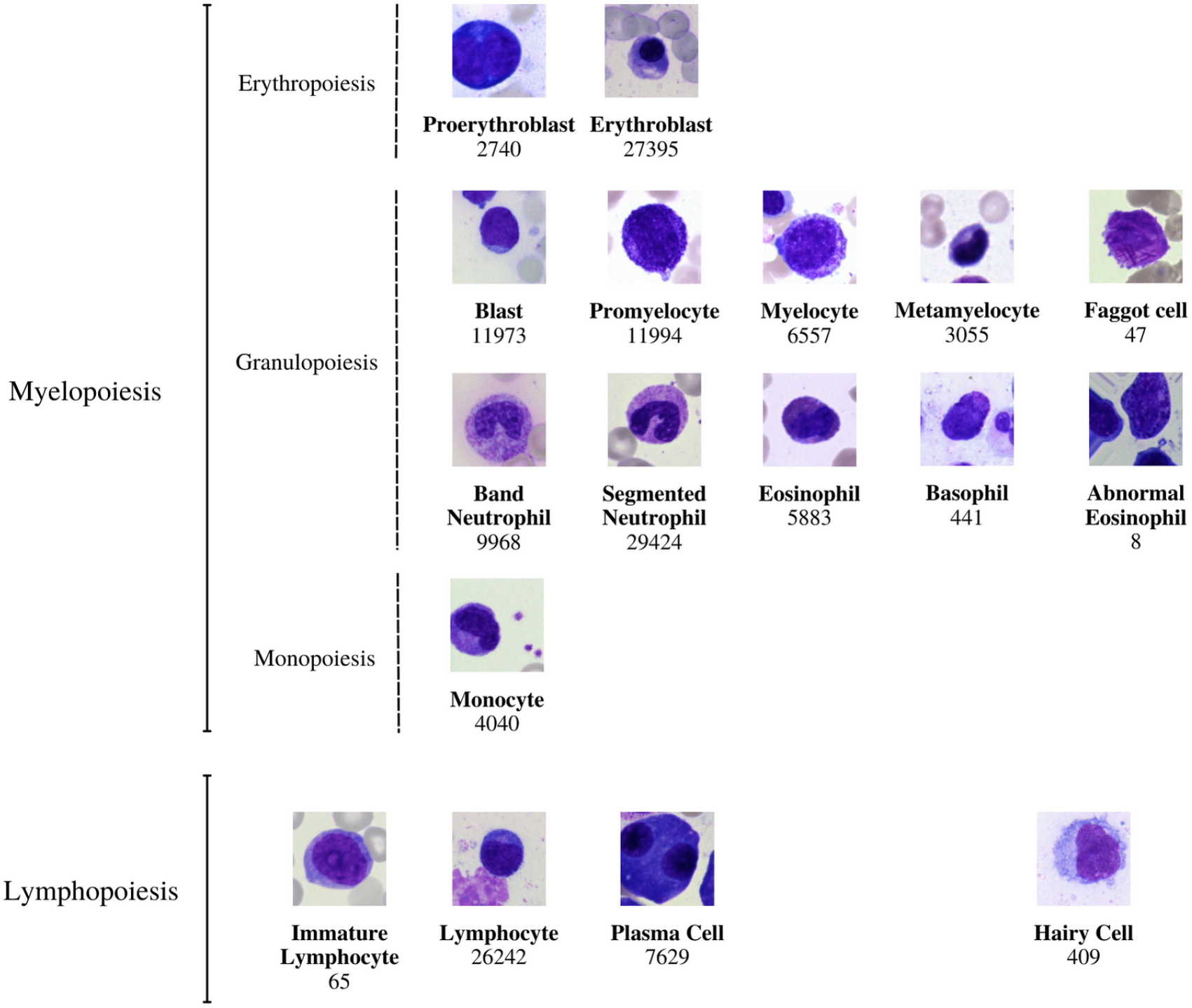
The 21 morphological classifications of BM cells employed in this investigation have a similar structure. The classes are arranged into hematopoietic lineages in the following order: In accordance with standard practice, the main physiological classes of myelopoiesis and lymphopoiesis, as well as typical pathological classes and, are all included in the classification. As described in further detail in the main text, all cells were stained with the May-Grunwald-Giemsa/Pappenheim stain and photographed at a magnification of 340.

**Figure 2:**
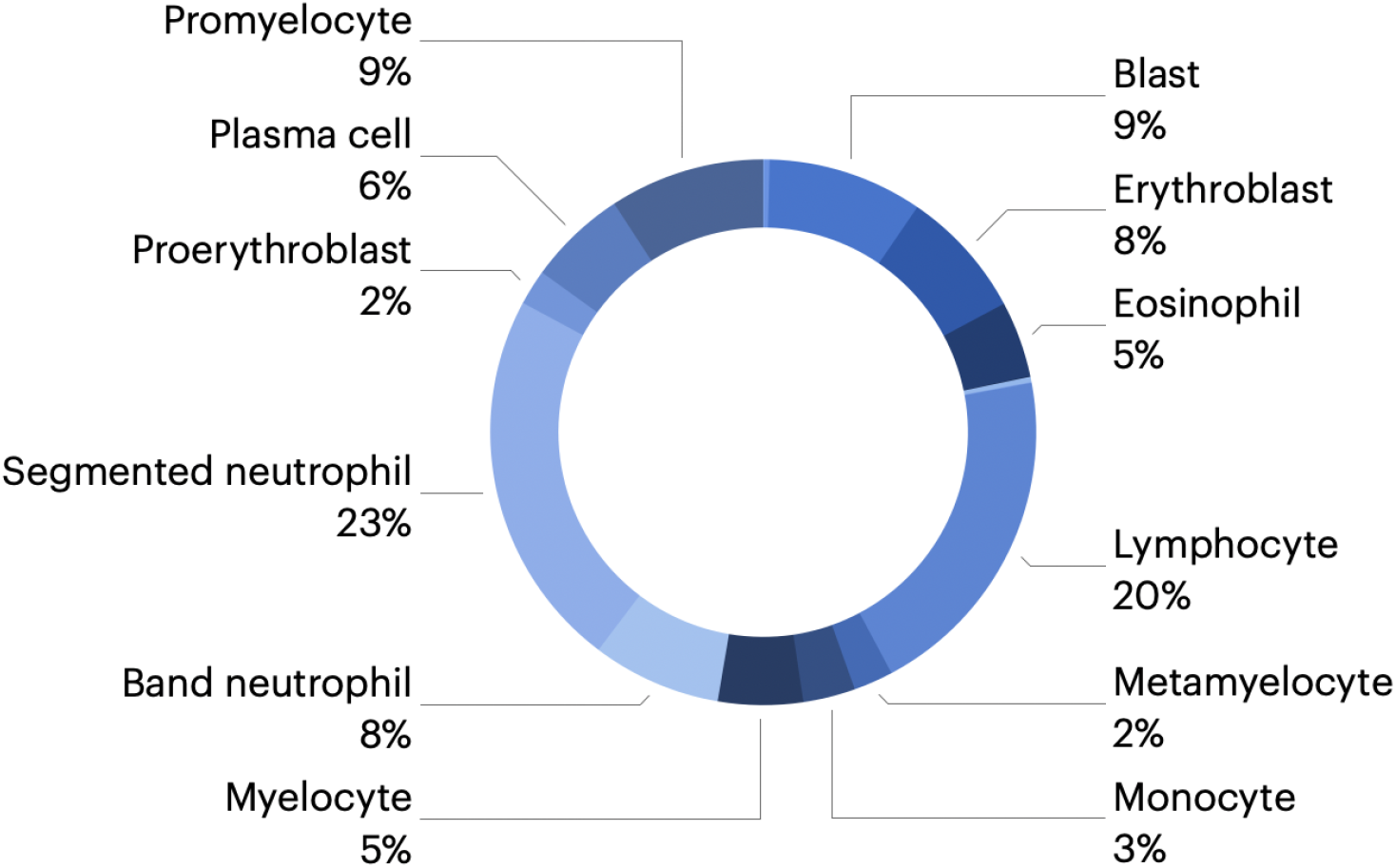
The pie chart shows the data distribution, we divided up the 171 374 nonoverlapping photos into the several classifications that were utilized.

### 3.2 Preprocessing

In this study we performed data augmentation in order to rectify class imbalances. Data augmentation is a technique used to generate new data from a set of existing data. In the case of images, new data can be created by applying a variety of transformations to an image. Some such transformations are rotations, translations, zooming in or out, noise insertion, cropping, and flipping horizontally or vertically.

Data augmentation is especially effective when used to address uneven distribution of class data. In our case, some of the classes had as few as 8 images, while others had as many as 29000. Augmenting the data helped address this issue, increasing the number of images we had for classes that were under-represented. This reduces bias towards over-represented classes.

We augmented the under-represented classes to roughly 20,000 images per class. The random transformations we performed included rotating between 0° to 90°, zooming in/out, flipping horizontally and/or vertically, and translating. Examples of these augmented images are shown in Figure 3.

**Figure 3:**
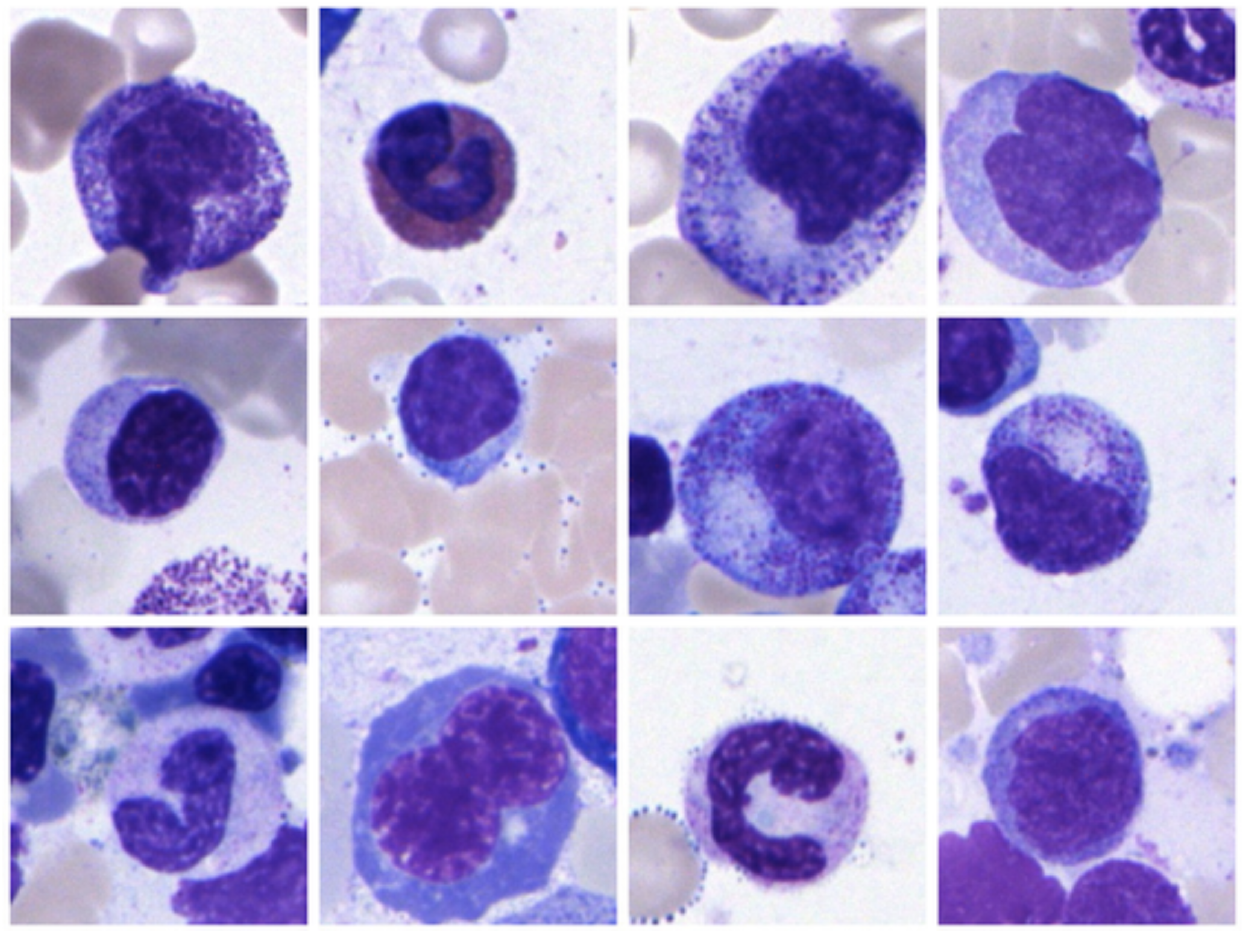
The augmented data from each classes are shown in the figure. The random transformations we performed included rotating between 0° to 90°, zooming in/out, flipping horizontally and/or vertically, and translating

**Figure 4:**
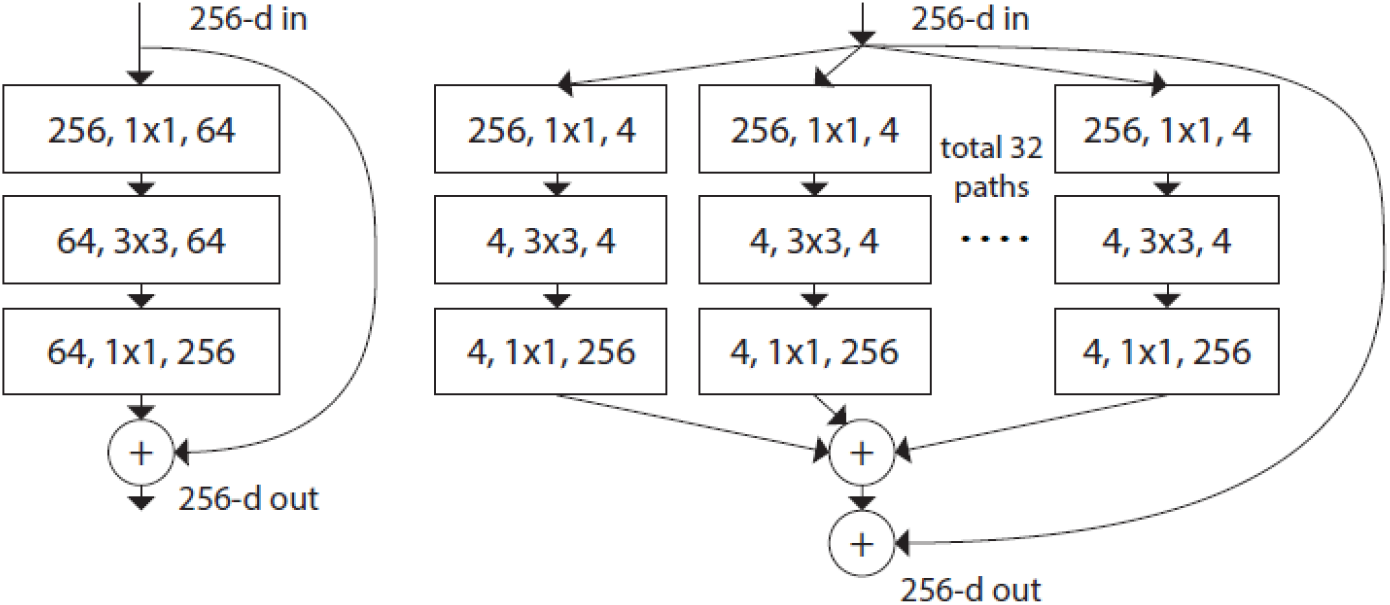
ResNet [56] is shown on the left. A ResNeXt block with cardinality = 32 and nearly the same complexity as the previous block is shown on the right. A layer is represented as (number of in channels, filter size, number of out channels)..

**Figure 5:**
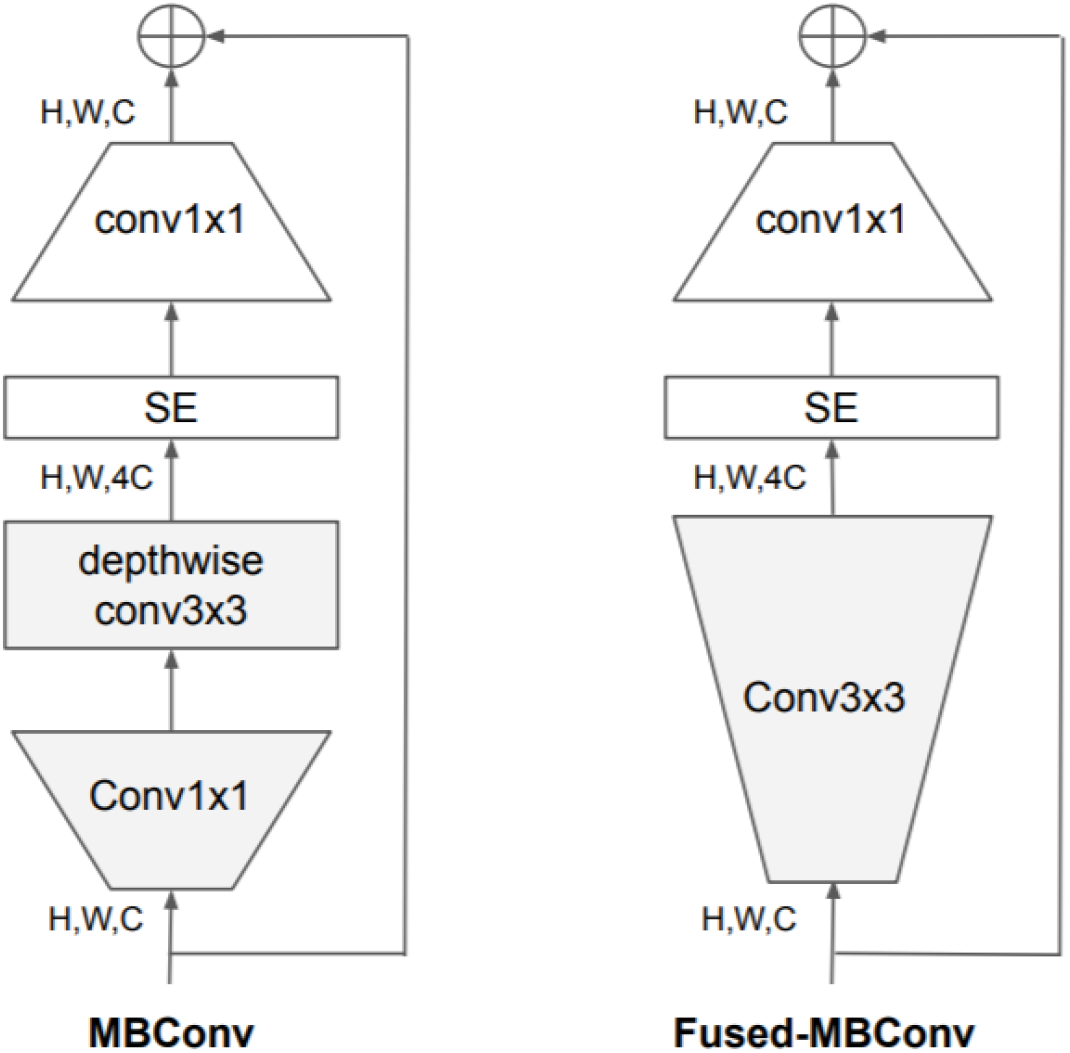
The MBConv and Fused-MBConv blocks are shown in the diagram above.

We also removed images that were irrelevant to our study. Images classified as ‘Artefacts’, ‘Other’, and ‘Unidentifiable’ were discarded before the augmentation process.

### 3.3 Models Implemented

As a gold standard, we implemented EfficientNetV2 [52] and ResNexT50 models [53, 54], while our main model for the pipeline is CoAtNet [55]. We chose the CoAtNet because of it’s Convolution and Attention based model architecture.

#### 3.3.1 ResNexT50

In the ImageNet Large Scale Visual Recognition Challenge 2016 competition, we employed the ResNeXt-50 architecture built by Xie et al, a successful image classification network that came in second place. 36 Peripheral blood smears have previously been classified using a network-like topology,14 making it an obvious candidate for BM cell classification. The limited amount of hyperparameters in the ResNet architecture is one of its advantages. The base architecture with 32 cardinalities and a 4d bottleneck width built ResNeXt-50. For ease of reference, this network is referred to as ResNeXt-50 (32×4d). It should be noted that the template’s input/output width is set to 256-d and that the feature map’s subsampled widths are always doubled.

Neurons in artificial neural networks perform inner product which can be thought of as a form of aggregating transformation:

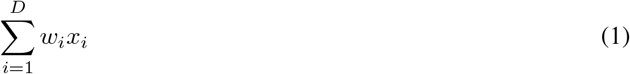

where *x* = [*x*_1_, *x*_2_, …, *x*_*D*_] is a D-channel input vector to the neuron and *w*_*i*_ is a filter’s weight for the *i*-th channel. This is the elementary transformation that’s done by the convolutional and fully-connected layers.

Aggregated transformations are presented as:

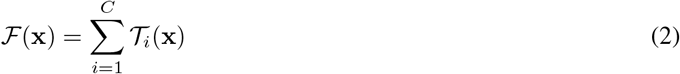

where *T*_*i*_(*x*) can be an arbitrary function. Analogous to a simple neuron, *T*_*i*_ should project x into an embedding and then transform it. The cardinality of the network, *C*, is the size of the set of transformations that will be aggregated can be an arbitrary number. While the dimension of width is related to the number of simple transformations, the dimension of cardinality controls the number of more complex transformations.

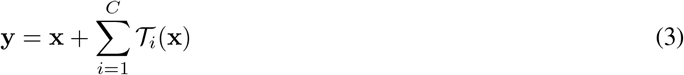

#### 3.3.2 EfficientNetV2

EfficientNetV2 is a new family of convolutional networks with quicker training speeds and higher parameter economy. These models are developed using a mix of neural architecture search and scaling, which are both designed to optimize training time and parameter efficiency. The EfficientNetV2 backbone differs significantly from the original EfficientNet in many key ways: MBConv and the new fused-MBConv are utilized extensively in the early layers of EfficientNetV2[52]. It also likes a lower MBConv expansion ratio since smaller expansion ratios often have less memory access overhead. In order to compensate for the decreased receptive field caused by the smaller 3×3 kernel size, EfficientNetV2 adds extra layers. Because of its huge parameters and memory access cost, it is possible that EfficientNetV2 totally eliminates stride-1 in the original EfficientNet.

EfficientNetV1 used MBConv layers with depth-wise convolutions in its design. However, current accelerators typically cannot fully use the reduced parameters and FLOPs of depthwise convolutions. As a result, reducing FLOPs doesn’t always translate into faster training. EfficientNetV2 uses MBConv and Fused-MBConv instead of depthwise convolution. The MBConv and Fused-MBConv blocks are shown in the figuer. In both blocks, an 1×1 convolution is applied to the SE module. The MBConv block employs an 1×1 conv and a depthwise convolution with a 3×3 filter in the early stages. A 3×3 filter is applied to a single convolution layer created by fusing 1×1 and 3×3 convolution together in the Fused-MBConv block.

#### 3.3.3 CoAtNet

CoAtNet is a hybrid model that merges Convolutional Neural Networks and Transformer models. CoAtNet was developed because it combines the capabilities of both ConvNet and Transformer into a single network CoAtNe delivers cutting-edge performance with varying data volumes while using the same resource. Inductive biases in CoAtNet allow it to generalize like ConvNets. In addition, CoAtNet benefits from better transformer scalability and quicker convergence, increasing its efficiency.

Generalization and model capacity are the two main elements from which hybridizing convolution and attention in machine learning is examined. The research demonstrates that convolutional layers have greater generalization while attention has higher model capacity. We can get greater generalization and capacity by merging convolutional and attention layers.

This hybrid model is more focused on image classification and is based on two key features: (a) Depthwise convolution and self-attention may readily be combined using simple relative attention. (b) By stacking convolution layers and attention layers in a logical way, generalization, capacity, and efficiency are greatly increased.

The model architecture, shown in Figure 6, consist of convolution and self-attention operations. The convolutional layer reduces the dimensionality of the input. The MBconv blocks are a type of image residual block that has an inverted structure. The first MBconv block expands the input by 4x before performing a depthwise convolution to capture the spatial interaction and the second block will compress it before adding a residual. Depthwise convolution performs convolution for each channel separately can be expressed as:

**Figure 6:**
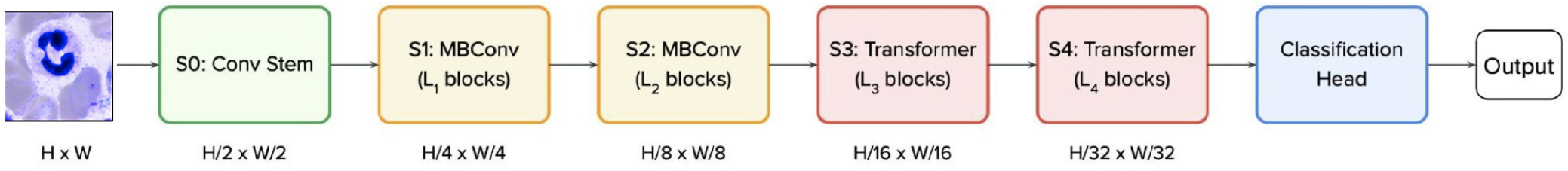
The CoAtNet architecture is shown in the figure. An input picture of size HxW, we use convolutions at the first stem stage (S0) to decrease the size to H/2 x W/2, which is the final size. With each step, the size of the object continues to shrink. The number of layers is denoted by the letter Ln. Then, the first two stages (S1 and S2) mostly use MBConv building pieces, which are composed of depth wise convolution operations. The last two stages (S3 and S4) are mostly comprised of Transformer blocks with a high degree of relative self-attention. In contrast to the preceding Transformer blocks in ViT, we employ pooling between stages in this block, which is comparable to the Funnel Transformer block. We then use a classification head to provide predictions.

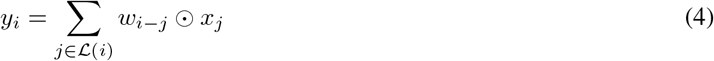

where *x*_*i*_, *y*_*i*_ ∈ ℝ^*D*^ are the input and output at position *i* respectively, and (*i*) denotes a local neighborhood of *i*, e.g., a 3 ×3 grid centered at *i* in image processing. For the self-attention blocks and feed-forward network (FNN) module uses the same expansion-compression structure, similar to the MBConv blocks. In comparison, self-attention allows the receptive field to be the entire spatial locations and computes the weights based on the re-normalized pairwise similarity between the pair (*x*_*i*_, *x*_*j*_) :^2^

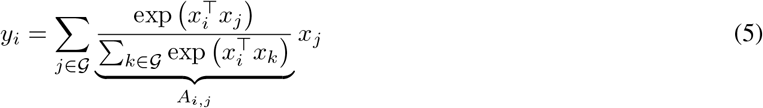

where 𝒢 indicates the global spatial space. The last three stages of a CoAtNet can be either a Convolution or a Transformer block, resulting in multiple combinations for the model architecture. Table 1 displays the family of CoAtNet models that have different sizes, number of blocks and hidden channels in the model architecture.

**Table 1:** L specifies the number of blocks, and D denotes the number of channels in the hidden dimension. We always utilize the kernel size 3 for all Conv and MBConv blocks, no matter what. Following [22], we increase the size of each attention head in all Transformer blocks to 32. The expansion rate for the inverted bottleneck is always 4, while the expansion (shrink) rate for the SE is always 0.25. The inverted bottleneck is also known as the inverted bottleneck.

| Stage | Size | CoAtNet-0 |  | CoAtNet-1 |  | CoAtNet-2 |  | CoAtNet-3 |  | CoAtNet-4 |  |
| --- | --- | --- | --- | --- | --- | --- | --- | --- | --- | --- | --- |
| S0-Conv | 1/2 | L=2 | D=64 | L=2 | D=64 | L=2 | D=128 | L=2 | D=192 | L=2 | D=192 |
| S1-MbConv | 1/4 | L=2 | D=96 | L=2 | D=96 | L=2 | D=128 | L=2 | D=192 | L=2 | D=192 |
| S2-MBConv | 1/8 | L=3 | D=192 | L=6 | D=192 | L=6 | D=256 | L=6 | D=348 | L=12 | D=348 |
| S3-TFMRel | 1/16 | L=5 | D=398 | L=14 | D=398 | L=14 | D=512 | L=14 | D=768 | L=28 | D=768 |
| S4-TFMRel | 1/32 | L=2 | D=768 | L=2 | D=768 | L=2 | D=1024 | L=2 | D=1536 | L=2 | D=1536 |

Input-Adaptive Weighting makes self-attention more prone to capture the relationships between different elements in the input and Global Receptive Field is the larger receptive field that’s used in self attention. An optimal model architecture involves Input-Adaptive Weighting and Global Receptive Field characteristics of self-attention and the Translation Equivariance that is featured in CNNs as a way to improve generalization for a limited size dataset. The overall idea is to sum a global static convolution kernel with the adaptive attention matrix, after or before the softmax initialization:

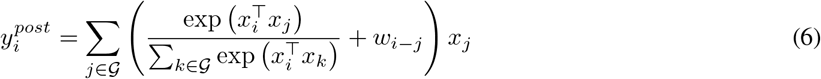

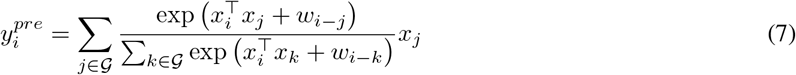

### 3.4 Evaluation Metrics

Most morphological classifications were accurately predicted by our trained model. Because neural networks are data-driven learning algorithms, their classification performance improves as the number of training sample data increases. Precision and recall, which are commonly used measurements of accuracy, precision, and recall, were utilized to evaluate our training method.

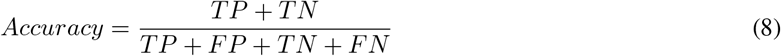

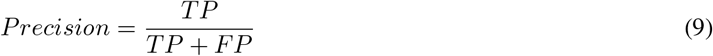

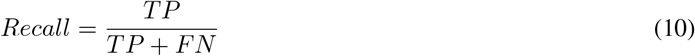

A class’s “true positives (TP)” and “true negatives (TN)” are determined by counting the number of predictions that agree with the ground truth in each category. “False positives (FP)” and “false negatives (FN)”, on the other hand, represent the amount of predictions that are categorized or not classified into a specific class despite the fact that the ground truth contradicts their classification.

The accuracy is calculated by dividing the number of properly identified samples by the total number of samples in the assessment data set. This measure is one of the most often utilized in ML applications in medicine, but it is also renowned for being deceptive in the situation of various class proportions, since just assigning all samples to the predominant class is a simple method to get high accuracy.

Recall determines the ratio of properly categorized positive samples to all samples allocated to the positive class, also known as the sensitivity or the True Positive Rate (TPR).

The precision reflects the fraction of the recovered samples that are relevant and is determined as the ratio of properly categorized samples to all samples allocated to that class. When the costs of False Positives are substantial, precision is a suitable metric to use.

## 4 Results and Discussion

The use of neural networks has been demonstrated to be effective in a variety of image classification challenges [57]. Using BM smears from a large patient cohort, we offer a comprehensive annotated high-quality data set of microscopic images collected from BM smears that may be utilized as a reference for developing machine learning algorithms for morphological categorization of diagnostically significant leukocytes. To the best of our knowledge, this picture database is the most comprehensive one currently accessible in the literature in terms of the number of patients, diagnoses, and single-cell images that are included within it. We utilized the data set to train and evaluate a novel convolutional and attention network-based model for cytology morphological classification, which was then compared with existing state-of-the-art CNN models.

The accurate distinction of various morphological classes, as previously stated, may be challenging, particularly when they are strongly tied to the leukocyte differentiation lineage; however, this is not always the case. Consequently, certain predictions of the network may be regarded as reasonable, even if they deviate from the ground truth by the human annotator, because of the inherent uncertainty associated with the morphological categorization. In the case of segmented neutrophils and band neutrophils, which are subsequent morphological classes in the ongoing process of myelopoiesis, misunderstanding between the two might be deemed tolerated in certain circumstances [58].

As shown in Table 2, the accuracy, precision, and recall values achieved by the CoAtNet (C), ResNext2 (R), and EfficientNetV2 (E) for each of the distinct morphological classes. Our proposed CoAtNet model outperformed both of the other models because of its attention network property that increased the learning curve for the algorithm which is represented using a precision-recall curve in figure 7.

**Table 2:** Comparison of Multi-Class Classification Metrics Results amongst Various Models Used. The C, R, and E represents CoAtNet [55], ResNexT50 [53, 54], and EffcientNetV2 [52] respectively.

| Class Name | Accuracy |  |  | Precision |  |  | Recall |  |  |
| --- | --- | --- | --- | --- | --- | --- | --- | --- | --- |
|  | C | R | E | C | R | E | C | R | E |
| Band neutrophils | **0.96** | 0.89 | 0.85 | **0.97** | 0.84 | 0.78 | **0.96** | 0.87 | 0.75 |
| Segmented neutrophils | **0.97** | 0.91 | 0.89 | **0.95** | 0.89 | 0.86 | **0.97** | 0.85 | 0.88 |
| Lymphocytes | **0.91** | 0.82 | 0.81 | **0.94** | 0.83 | 0.84 | **0.93** | 0.78 | 0.81 |
| Monocytes | **0.77** | 0.59 | 0.47 | **0.81** | 0.62 | 0.56 | **0.79** | 0.63 | 0.57 |
| Eosinophils | **0.85** | 0.68 | 0.64 | **0.91** | 0.65 | 0.62 | **0.88** | 0.69 | 0.6 |
| Basophils | **0.64** | 0.27 | 0.14 | **0.74** | 0.11 | 0.15 | **0.7** | 0.17 | 0.15 |
| Metamyelocytes | **0.88** | 0.21 | 0.29 | **0.91** | 0.15 | 0.21 | **0.89** | 0.12 | 0.24 |
| Myelocytes | **0.85** | 0.75 | 0.72 | **0.88** | 0.65 | 0.69 | **0.87** | 0.59 | 0.66 |
| Promyelocytes | **0.97** | 0.89 | 0.86 | **0.98** | 0.79 | 0.8 | **0.98** | 0.73 | 0.78 |
| Blasts | **0.96** | 0.91 | 0.92 | **0.94** | 0.71 | 0.84 | **0.98** | 0.88 | 0.8 |
| Plasma cells | **0.94** | 0.89 | 0.87 | **0.93** | 0.74 | 0.85 | **0.95** | 0.75 | 0.82 |
| Proerythroblasts | **0.84** | 0.68 | 0.57 | **0.89** | 0.53 | 0.58 | **0.85** | 0.5 | 0.55 |
| Erythroblasts | **0.98** | 0.87 | 0.88 | **0.99** | 0.78 | 0.86 | **0.98** | 0.76 | 0.83 |
| Hairy cells | **0.93** | 0.51 | 0.45 | **0.92** | 0.46 | 0.4 | **0.88** | 0.41 | 0.42 |
| Abnormal eosinophils | **0.43** | 0.18 | 0.12 | **0.42** | 0.11 | 0.09 | **0.39** | 0.13 | 0.14 |
| Immature lymphocytes | **0.63** | 0.22 | 0.16 | **0.65** | 0.13 | 0.15 | **0.66** | 0.12 | 0.16 |
| Faggot cells | **0.77** | 0.23 | 0.19 | **0.83** | 0.16 | 0.1 | **0.87** | 0.18 | 0.11 |

**Figure 7:**
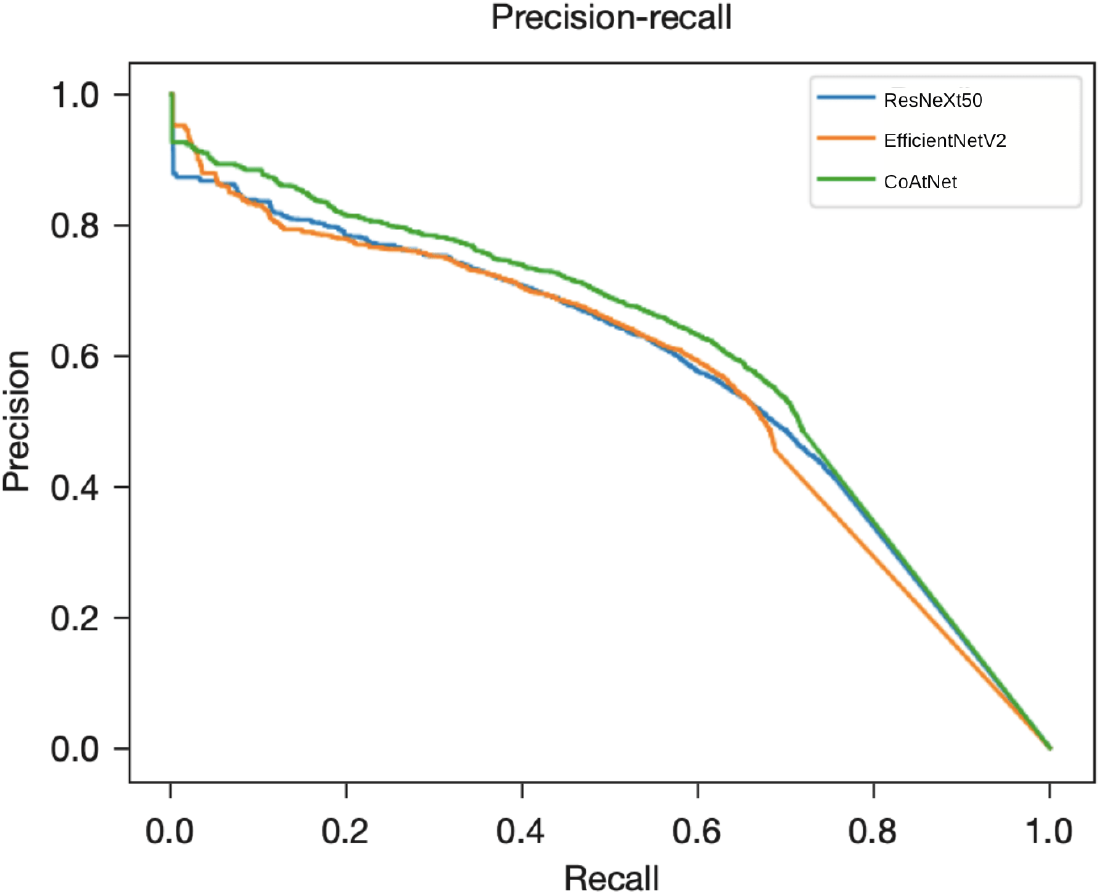
The figure shows precision-recall curve for each of the models we trained. the The trade off between precision and recall may be seen in the precision-recall curve for various threshold values. A low false negative and false positive rate is associated with a higher area under the curve..

For classes in which there are just a few training samples available, such as faggot cells or diseased eosinophils, the classifier performs less well, as would be anticipated for a data-driven strategy [59, 60]. There would be a greater need for training data if the image-classification job was focused on the detection of these particular cell types. It is also possible that training a binary classifier rather than a complete multiclass classifier will result in improved prediction performance.

Because neural networks are created in a data-driven manner based on the training set, the classification judgments made by neural networks do not lend themselves to straightforward human interpretation. However, in order to obtain insight into the classifications made by these algorithms, a number of explainability approaches have recently been created to aid in their investigation [61]. As part of our effort to determine which regions of the input images are important for the network’s classification decisions, we analyzed the CoAtNet model using SmoothGrad[62] and Grad-CAM[63], two recently developed algorithms that have been shown to meet the fundamental requirements for explainability methods (ie, sensitivity to data and model parameter randomization) [64]. As seen in Figure 8, the model has trained to focus on the important input of a single-cell patch (ie, the primary leukocyte depicted in it) while disregarding background characteristics such as erythrocytes, cell debris, and pieces of other cells visible in the patch. Furthermore, specific defining structures that are known to be relevant to human examiners when classifying cells appear to play a role in the network’s attention pattern, such as the cytoplasm of eosinophils and the cell membrane of hairy cells, which appear to play a role in the network’s attention pattern. Although these analyses, when used as post classification explanations, do not in and of themselves guarantee the correctness of a particular classification decision, they can help to increase confidence that the network has learned to focus on relevant features of the single-cell images and that predictions are based on reasonable characteristics.

**Figure 8:**
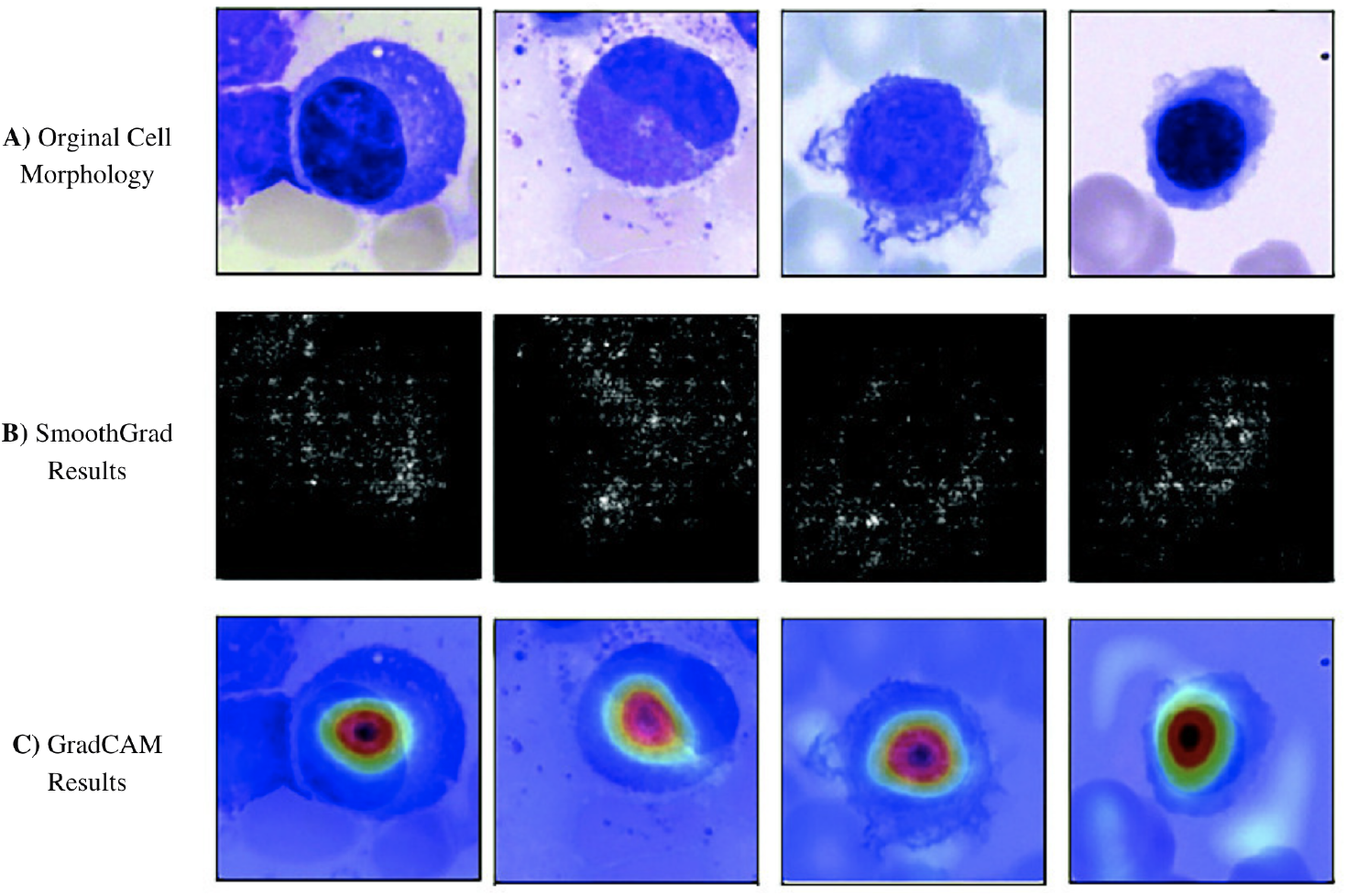
Original photos classified properly by the network are presented in the top row. All cells were stained with the May-Gruenwald-Giemsa/Pappenheim stain and photographed at 340 magnification, as described in the primary text. The center row displays analysis using the SmoothGrad algorithm. The brighter a pixel looks, the more it contributes to the categorization judgment made by the network. Results of a second network analysis approach, the Grad-CAM algorithm, are exhibited in the bottom row as a heat map superimposed on the original picture. The areas of the image that contain essential information are highlighted in red. Both analytical approaches imply that the network has trained to concentrate on the leukocyte while disregarding background structure. Note the network’s focus on traits known to be significant for certain cell types, such as the cytoplasmic organization of eosinophils or the nuclear architecture of plasma cells.

As a whole, the results are positive and promising, with good accuracy and recall values achieved for the vast majority of diagnostically relevant classes studied. Our results are consistent with previous findings in other areas of medical imaging, where attention-based image classification tasks have outperformed approaches that rely on the extraction of image features to attain higher standards [65, 66, 67, 68]. The most important component of the successful use of CNNs is a training data set that is sufficiently big and of good quality [69].

## 5 Conclusion

As part of the current investigation, we mostly used a single-center strategy, with all BM smears included for training having been stained, captured, and processed in the same laboratory. The network presented in this paper performs well in such an environment, which is quite promising. Our proposed convolution and attention network model (CoAtNet) outperformed the current state-of-the-art CNN models in classifying cells and even took a huge leap in accurately classifying classes with low sample sizes. This shows how the attention network could be used in similar datasets in the future as well.

Future research may help to lessen the importance of label noise (eg, by using semi- or unsupervised methods as having been applied to processes such as erythrocyte assessment46 or cell cycle reconstruction). Increased growth of the morphological database, preferably as part of multi-centric research and using a variety of scanner hardware, will almost certainly improve the performance and resilience of the network, particularly for classes with a smaller size in the current data set.

However, because of the large number of cases and diagnoses included in our study, we anticipate that the data set will fairly accurately represent the morphological variation seen in most cell groups. The purpose of this research is to evaluate the morphology of the adult BM. It would be fascinating to expand the study to include samples from newborns and young children, particularly for lymphoid cells. The performance of our network in a real-world diagnostic environment will need more investigation. The range of diagnostic modalities employed in hematology suggests that the integration of supplementary data (for example, from flow cytometry or molecular genetics) would improve the quality of predictions made by neural networks.

## Data Availability

All data produced are available online at

https://wiki.cancerimagingarchive.net/pages/viewpage.action?pageId=101941770#101941770171ba531fc374829b21d3647e95f532c

## Notes

### Competing Interest Statement

The authors have declared no competing interest.

### Funding Statement

This study did not receive any funding

